# The effects of white matter hyperintensities on MEG power spectra in cognitively healthy aging

**DOI:** 10.1101/2024.05.15.24307438

**Authors:** Lucia Torres-Simón, Pablo Cuesta, Alberto del Cerro, Sandra Doval, Brenda Chino, Lucía Hernández, Elizabeth B. Marsh, Fernando Maestú

**Author notes:** Corresponding author: Lucia Torres-Simon. (a) Center of Cognitive and Computational Neuroscience (UCM), (b) Department of Experimental Psychology, Cognitive Processes and Speech Therapy, Universidad Complutense de Madrid (UCM), 28223 Pozuelo de Alarcón, Madrid, Spain. **Authors’ s contributions** **Lucia Torres-Simon:** Conceptualization, Methodology, Formal Analysis, Investigation, Writing-original Draft, Writing - Review & Editing, Visualization, Project administration, Funding acquisition. **Pablo Cuesta:** Conceptualization, Methodology, Formal Analysis, Investigation, Writing-original Draft, supervision and Funding acquisition. **Alberto del Cerro:** Methodology, Formal Analysis, Investigation, Writing-original Draft, Funding acquisition. **Brenda Chino:** Writing - Review & Editing and Supervision. **Lucia Hernandez:** **Elisabeth B. Marsh:** Writing - Review & Editing, Supervision, Validation. **Miguel Yus:** **Fernando Maestú:** Writing - Review & Editing, Resources, Supervision and Funding acquisition.

## Abstract

**Objective:** This study sought to identify magnetoencephalography (MEG) power spectra patterns associated with cerebrovascular damage (white matter hyperintensities - WMH) and their relationship with cognitive performance and brain structure integrity in aging individuals without cognitive impairment.

**Methods:** We hypothesized a “slowness” pattern characterized by increased power in δ and θ bands and decreased power in the β band associated with the severity of vascular damage. MEG signals were analyzed in cognitively healthy older adults to investigate these associations.

**Results:** Contrary to expectations, we did not observe an increase in δ and θ power. However, we found a significant negative correlation between β band power and WMH volume. This β power reduction was linked to structural brain changes, such as larger lateral ventricles, reduced white matter volume, and decreased fractional anisotropy in critical white matter tracts, but not to cognitive performance. This suggests that β band power reduction may serve as an early marker of vascular damage before the onset of cognitive symptoms.

**Conclusion:** Our findings partially confirm our initial hypothesis by demonstrating a decrease in β band power with increased vascular damage but not the anticipated increase in slow band power. The lack of correlation between the βpow marker and cognitive performance suggests its potential utility in early identification of at-risk individuals for future cognitive impairment due to vascular origins. These results contribute to understanding the electrophysiological signatures of preclinical vascular damage and highlight the importance of MEG in detecting subtle brain changes associated with aging.

## Introduction

Besides from pure vascular Dementia (VaD) or major vascular cognitive impairment (VCI) which is responsible for up to 20% of cases of dementia (Kalaria, 2018; O’Brien and Thomas, 2015; Rizzi et al., 2014), cerebrovascular diseases (CBVD) are major comorbid contributors to the progression of other neurodegenerative diseases; for example, it can be observed in 50%–90% of AD patients (Santos et al., 2017), and about 50% of dementias worldwide (Wardlaw et al., 2019). CBVD involves a variety of medical conditions or changes in the cerebral or systemic vasculature with brain impact, including a huge number of pathologies and etiologies.

Vascular dementia (VaD) or major vascular cognitive impairment (VCI) accounts for approximately 20% of all dementia cases (Kalaria, 2018; O’Brien & Thomas, 2015; Rizzi et al., 2014). Additionally, cerebrovascular diseases (CBVD) significantly contribute to the progression of other neurodegenerative diseases, with evidence of their presence in 50%–90% of Alzheimer’s disease (AD) patients (Santos et al., 2017) and an implication in about half of the global dementia cases (Wardlaw et al., 2019). CBVD represents a spectrum of conditions affecting both cerebral and systemic vasculature with neurological implications, which include a diverse array of pathologies and etiologies. Given this diversity, it is highly challenging to identify electrophysiological signatures that are universally applicable across these conditions. Consequently, research efforts may benefit from a narrowed focus on specific cerebrovascular damage rather than broad diagnostic groups.

Within the spectrum of vascular pathologies, white matter hyperintensities (WMHs) are a prominent feature. These are observable consequences of blood-brain barrier degradation secondary to small vessel disease (SVD) and are detectable via neuroimaging (Regenhardt et al., 2018). WMHs are not exclusive to dementia patients with VCI or AD (Jellinger, 2013), but are also prevalent in the healthy elderly population (Debette & Markus, 2010; Kloppenborg et al., 2014), with their prevalence escalating with advancing age (Alber et al., 2019; Van Leijsen et al., 2017). The clinical significance of WMHs is underscored by their association with an increased risk of dementia of all types and their substantial contribution to mixed dementia. Furthermore, WMHs are implicated in about one-fifth of strokes worldwide (Chutinet & Rost, 2014; Debette & Markus, 2010; Norrving, 2008; Pantoni, 2010).

The presence and progression of WMHs in the brain have been associated with general cognitive decline, particularly affecting attention, and executive function. This is evident in both healthy older populations (Arvanitakis, Fleischman, et al., 2016; Kloppenborg et al., 2014), and dementia patients (Arvanitakis, Capuano, et al., 2016; Lam et al., 2021; Mortamais et al., 2014; Prins & Scheltens, 2015; Van Den Berg et al., 2018). Moreover, WMHs have been extensively linked to structural and pathophysiological changes in the brain, including grey matter atrophy (Gaubert et al., 2021; Jang et al., 2017), cortical thinning (Duering et al., 2012), ventricular enlargement (Bjerke et al., 2014), reduced hippocampal volume (Porcu et al., 2020), and compromised white matter integrity as studied with diffusion tensor imaging (DTI) (Veldsman et al., 2020).

Given their high incidence and their substantial contribution to cognitive and brain impairment—along with the potential for treatment—WMHs hold particular significance for the early differentiation of the pathophysiological origins of cognitive decline in the elderly. This could lead to more accurate prognoses and the development of new therapeutic targets aimed at slowing the progression of dementia.

Electrophysiological neuroimaging techniques hold particular significance for the early differentiation of cognitive disorders, as they yield valuable insights into brain function and network dynamics that may elude standard structural imaging methods and cognitive assessments. These neurophysiological modalities, distinct from other functional neuroimaging approaches, capture a broad spectrum of brain oscillations ranging from delta to gamma frequencies. They directly measure neural activity, in contrast to the indirect assessments of blood flow or metabolism utilized by alternative techniques. Furthermore, they permit repeated measurements without posing any risk to patients. Among the array of electrophysiological tools, electroencephalography (EEG) and magnetoencephalography (MEG) are prominent. EEG, while being more cost-effective and widely used in clinical settings, is complemented by MEG. MEG offers superior spatial resolution for neural source estimation and a higher signal-to-noise ratio, particularly within the higher frequency bands. Additionally, MEG is considered more patient-friendly and has been integrated into clinical practice in some hospitals (Hoshi et al., 2022).

Research on electrophysiological spectral signatures via magnetoencephalography (MEG) has been pivotal for early detection in neurodegenerative diseases, notably Alzheimer’s disease and mild cognitive impairment (López-Sanz et al., 2016, 2018, 2019; Nakamura et al., 2018). However, spectral analysis using MEG in vascular cognitive impairment (VCI) is sparse, with limited EEG studies identified in our systematic review (Torres-Simon et al., 2022).

In the reviewed EEG studies, a common dementia profile emerged in VCI patients, characterized by increased delta and theta power and reduced beta power, indicative of cerebrovascular disease severity (Al-Qazzaz, Ali, et al., 2017; Al-Qazzaz, Hamid Bin Mohd Ali, et al., 2017; Babiloni, Binetti, et al., 2004; Moretti et al., 2004; Neto et al., 2015; Schreiter Gasser et al., 2008; van Straaten et al., 2012; Wu et al., 2014). This pattern was more pronounced in VCI than AD. Alpha band analysis showed greater variability, likely due to differing frequency band definitions. These EEG studies focused on diagnosed mild or major VCI cases, involving more cognitive impairment and heterogeneity than our sample. They compared groups (VCI vs. HC or AD), while our goal was to correlate cerebrovascular damage with power spectra patterns directly. In this concern, another paper out of the scope of the described review was published in 2020, including 35 cognitively healthy patients with and without WMH (Quandt et al., 2020). In this study, they performed correlation analysis between WMH volume and EEG resting-state signal, and again they found higher theta power related to greater WMH volume. Keeping all these conditions in mind we built up some exploratory hypothesis looking at previous knowledge:

H1: We expect to find a broad “slowness” pattern, specially, higher power in the δ, θ, and lower in β band associated with the severity of the vascular damage.

H1.1: The cluster emerged will not correlate with cognitive performance, as our participants are still cognitively healthy.

H1.2.: The cluster emerged will negatively correlate with brain structure (i.e., grey, and white matter integrity), as it may be affected even before cognitive symptoms are evidenced.

## Methods

### Participants

Participants were recruited from three Spanish national projects (PSI2009-14415-C03-01, PSI2012-38375-C03-01, PSI2015-68793-C3-1-R) on dementia research and early detection. Data collection spanned 2010-2018 at three Madrid clinical centers: “Hospital Universitario Clinico San Carlos,” the “Center for Prevention of Cognitive Impairment,” and the “Seniors Center of Chamartín District.” The protocol included MEG recordings, MR imaging (T1, FLAIR, DWI), neuropsychological tests, and genetic analysis. From 520 native Spanish-speaking participants, the following assessments were used for enrollment: MMSE (Lobo et al., 1979), Global Deterioration Scale (Reisberg et al., 1982), GDS-Short Form (Yesavage et al., 1982), FAQ (Pfeffer et al., 1982), and Lawton & Brody’s IADL questionnaire (1969). Exclusion criteria were psychiatric/neurological history and psychoactive or chronic medication use. Additional tests ruled out other cognitive decline causes. Informed consent was obtained; the study was approved by the ethics committee of “Hospital Universitario Clinico San Carlos.”

The original sample (N = 520) was refined to ensure data quality and reproducibility. After exclusions for age (<50 years), low MMSE (<26), radiological abnormalities (excluding WMH), incomplete 3D MRI or segmentation, absent or noisy MEG recordings, and WMH volume >3 SD above the mean, 220 were excluded, resulting in 300 cognitively healthy participants (Age = 66 ± 8; 68% females).

### Sample sub-groups according to a WMH volume cut point

Based on the established cut-off, we categorized participants into low vascular impairment (Low-VI) with WMH volumes below 1000 mm^3^, and mild vascular impairment (Mild-VI) above this threshold. Low-VI was identified in 59% of subjects (176 participants), whereas 41% (124 participants) exhibited Mild-VI, with WMH volumes reaching up to 23203 mm^3^. Subsequent analyses will be reported for the entire cohort and each subgroup separately, to examine how varying levels of vascular damage impact brain function, structure, and cognition in cognitively healthy older adults.

### Neuropsychological assessment

All patients underwent an exhaustive neuropsychological assessment to generate detailed cognitive profiles including: Direct and Inverse Digit Span Test (Wechsler Memory Scale, WMS-III; Wechsler, 1997), Immediate and Delayed Recall (WMS-III; Wechsler, 1997), Phonemic and Semantic Fluency (Controlled oral Word Association Test, COWAT; Benton and Hamsher, 1989), and Trail Making Test A and B (TMTA and TMTB; Reitan, 1958).

### Magnetic resonance recordings and analysis

T1-weighted and T2-weighted 3D FLAIR and DWI MRI sequences were available for each subject. These images were acquired in a General Electric 1.5 Tesla magnetic resonance scanner in the Hospital Universitario Clinico San Carlos, using a high-resolution antenna and a homogenization PURE filter (Fast Spoiled Gradient Echo sequence). The following acquisition parameters were followed for:

- The T1-weighted imaging: repetition time (TR) = 11.2 ms, echo time (TE) = 4.2 ms, inversion time (TI) = 450 ms, Field of View (FOV) = 25 cm, flip angle (FA) = 12°, 252 coronal slices (in-plain resolution: 256×256), voxel size: 0.98 × 0.98 × 1 mm3 and acquisition time ≃ 8:00 min.
- The T2-weighted 3D FLAIR: TR = 7000 ms, TE = 101 ms, TI = 2112 ms, FOV = 24 cm, 252 coronal slices (in-plain resolution: 256×112), voxel size: 0.94 × 0.94 × 1.6 mm3 and acquisition time ≃ 4:57 min.
- The DWI: TE/TR 96.1/12,000 ms; NEX 3 for increasing the SNR; 2.4 mm slice thickness, 128 × 128 matrix, and 30.7 cm FOV yielding an isotropic voxel of 2.4 mm; 1 image with no diffusion sensitization (i.e., T2-weighted b0 images); and 25 DWI (b = 900 s/mm2). Data were recorded with a single shot echo planar imaging sequence.

#### A) White matter hyperintensities segmentation (FLAIR and T1)

WMH segmentation (T1 and Flair) and lesion volume calculation was performed using two automatic computerized algorithms in the Lesion Segmentation Tool (LST) package included in the SPM12 toolbox as this combination was the most accurate compared with clinician’s manual segmentations (Torres-Simon et al., 2023). Lesion prediction algorithm (LPA) for the main calculation and lesion growth algorithm (LGA) to control the possible errors. We used the initial κ threshold with default value 0.3 as recommended by the developers (Schmidt et al., 2012), and subsequently the probabilistic threshold of 0.5.

#### B) Brain grey and white matter integrity (T1 and DWI)

The resulting images from the T1 sequence were processed using the Freesurfer software (version 6.1.0) and its specialized tool for automated cortical parcellation and subcortical segmentation (Fischl et al., 2002). The measures that were included in further analyses were total volumes (mm3) of gray matter, total white matter, lateral ventricles, and hippocampus; and the cortical thickness (mm). The volumes of bilateral structures were collapsed to obtain a single measure for each region.

DWI images were processed using probabilistic fiber tractography run in the automated tool *AutoPtx* (https://fsl.fmrib.ox.ac.uk/fsl/fslwiki/AutoPtx). The methodology is detailed in (Verdejo-Román et al., 2018) but we will briefly summarize the procedure followed to obtain the fractional anisotropy (FA) markers.

The DTI images were processed using FMRIB-FSL (Jenkinson et al., 2012), including adjustment for minor head motion, eddy-current artifacts, rotation of the gradient direction table in the same way than the images in the previous step, and non-brain tissue removal using the Brain Extraction tool of FSL (S. M. Smith, 2002). The calculation of FA maps was done by fitting the diffusion tensor using dtifit function (PREOBE and NFBC 1986) or the RESTORE method implemented in Camino (Generation R). Quality of the data was checked with automatic tools (i.e., DTIPrep tool; https://www.nitrc.org/projects/dtiprep/) and visual assessment. AutoPtx automated pipeline was used to run a probabilistic tractography of brainstem, projection, association, callosal, thalamic, and limbic fibers in each individual. Tracts were defined using seed, target, termination, and exclusion masks that were warped to the original diffusion space. The connectivity distributions were normalized and thresholded (M. De Groot et al., 2015), and the mean FA parameter per each tract was extracted. Bilateral tracts were collapsed to obtain a single measure. The pipeline ended up providing mean FA values for the 15 tracts of interest described in **¡Error! No se encuentra el origen de la referencia**..

### MEG data acquisition

Eyes-closed resting-state brain function was recorded for five minutes using a Vectorview system (Elekta Neuromag) at the Center for Biomedical Technology, Madrid. Participants were monitored for arousal via video and post-session conversation. MEG data was collected at a sampling frequency of 1000Hz and online band-pass filtered between 0.1 and 330 Hz. Each subject’s head shape was defined relative to three anatomical locations (nasion and bilateral preauricular points) using a 3D digitizer (Fastrak, Polhemus, VT, USA) and head motion was tracked through four head-position indicator (HPI) coils attached to the scalp. These HPI coils continuously monitored the subjects’ head movements, while eye movements were monitored by a vertical electrooculogram assembly (EOG) composed of a pair of bipolar electrodes. Raw recording data was first introduced to Maxfilter software (v 2.2, correlation threshold = 0.9, time window = 10 seconds) to remove external noise using the temporal extension of the signal space separation method with movement compensation (Taulu and Simola, 2006). Then, magnetometers data (Garcés et al., 2017) was automatically examined to detect ocular, muscle, and jump artifacts using Fieldtrip software (Oostenveld et al., 2011), which were visually confirmed by an MEG expert. The remaining artifact-free data was sectioned into four-second segments. Independent component analysis-based procedure (ICA) was applied to remove heart magnetic field artifacts and EOG components. Only those MEG recordings with at least 20 clean segments (80 seconds of brain activity) were used in further analysis. MEG clean time series were band-pass filtered (2 seconds padding) between 2 and 45Hz. Source reconstruction was carried out using a regular grid of 1 cm spacing in the Montreal Neurological Institute (MNI) template. The resulting model comprised 2459 sources homogeneously distributed across the brain. This model was linearly transformed to each subject’s space. The leadfield was calculated using a single shell model (Nolte, 2003). Sources time series were reconstructed using a Linearly Constrained Minimum Variance beamformer (Van Veen et al., 1997). Power spectrum of each grid’s node was computed by means of Fast Fourier Transform using Hanning tapers with 0.25 Hz smoothing. For each node, relative power was calculated by normalizing by total power over the 2 - to 30-Hz range. The source template with 2459 nodes in a 10 mm spacing grid was segmented into 78 regions of the Automated Anatomical Labeling atlas (AAL, Tzourio-Mazoyer et al., 2002), excluding the cerebellum, basal ganglia, thalamus and olfactory cortices. Those 78 regions of interest included 1202 of the original 2459 nodes. Trials were averaged across subjects ending up with a source-reconstructed power matrix of 1202 nodes x 113 frequency steps x 300 participants.

The power spectrum of each of the 1202 source locations were computed similarly to previous studies (de Frutos-Lucas et al., 2020; Garcés et al., 2013; Nakamura et al., 2018) with the Fieldtrip toolbox (Oostenveld et al., 2011) using a multitaper method (mtmfft) with discrete prolate spheroidal sequences (dpss) as windowing function and 1 Hz smoothing. The spectral power was calculated for the 2-30 Hz range in 0.25 Hz steps. For the analysis, we employed relative power that was calculated by normalizing each frequency step by the total power in the whole frequency range, ending up with a source-reconstructed power matrix of 1202 nodes x 113 frequency steps x 300 participants. Additionally, following previous studies of our group (D López-Sanz et al., 2016; María Eugenia López et al., 2020), we determined for each individual the individual alpha peak frequency (henceforth called Alpha peak) by visual inspection of the average power spectra of the occipital-parietal region of the brain. The bilateral AAL ROIs used for this purpose were: calcarine fissure and surrounding cortex; cuneus, lingual gyrus, right superior occipital lobe, middle occipital lobe, and inferior occipital lobe (María Eugenia López et al., 2020).

### Statistical procedure

This study seeks to identify resting-state MEG power spectra patterns linked to white matter hyperintensities (WMH) and their relationship with cognitive performance, and brain structure integrity in aging. We refrain from group comparisons, acknowledging the continuum of vascular load with age, which introduces high intragroup variability. Current literature suggests WMH volume data in healthy adults are inconsistent across studies due to sample and technical differences (Melazzini et al., 2021). Thus, we employ correlational analysis to meet our objective and determine a significant WMH volume threshold for cognitively healthy elderly.

The statistical pipeline was conceived to find brain regions (i.e., clusters) defined as a set of spatially adjacent nodes (sources locations) whose normalized power systematically (same sign and consecutive frequency steps) showed a significant correlation with the volume of WMH. The method relied on a cluster-based permutation test (CBPT) as implemented in Fieldtrip (Maris & Oostenveld, 2007; Oostenveld et al., 2011; Zalesky et al., 2010). The procedure was performed independently per three different samples: 1) the complete sample; 2) the Low VI subsample; and 3) the Mild VI subpopulation. The methodology involved the following steps:

1. Partial correlation tests between normalized power and WMH burden, controlling for age and brain volume, yielding Spearman’s rho and p-values per node/frequency.
2. Thresholding rho values at p < 0.01 (cluster-configuration threshold), creating matrices based on rho sign.
3. Clustering adjacent nodes in space-frequency using Matlab’s bwlabel function.
4. Clusters must exceed 1% of total nodes and show significance in three consecutive frequencies. This minimum size criterion was included to suppress spurious findings.
5. Rho values convert to Fisher’s Z, with cluster mass statistics computed as the sum of Z values.
6. Multiple comparison control is achieved by repeating steps 1-5, 5,000 times with shuffled data, creating a null distribution for family error rate control.
7. This empirical distribution allowed us to calculate the p value (henceforth called CBPT p value) for each original candidate cluster. Clusters with CBPT p < 0.05 are considered for further analysis as potential MEG markers.

In this framework, significant clusters act as functional units, whose representative MEG value is calculated by averaging the power of all the nodes and frequencies that make up the corresponding cluster. These MEG signatures (1 value per significant cluster and participant) were used in the following subsequent analysis.

a. Regardless of the population with which the cluster was obtained, we tested the Spearman correlation between the markers of each cluster and the volume of WMH in the 3 populations (complete, Low VI and Mild VI).
b. We carried out additional Spearman partial correlation analysis between the clusters’ signatures and indicators of brain structural and cognitive integrity.
c. Finally, we tested, for each cluster’s marker, the possible existence of between-subgroups (i.e., Low VI & Mild VI) differences by means of ANCOVA test with age and brain segmentation volume as covariates.

Statistical analyses were carried out using Matlab R2022b (The MathWorks Inc., Natick, MA, USA) and SPSS v. 22 (IBM, Armonk, NY, USA) software. All tests were two tailed, and the significance level threshold was set at p = 0.05 unless explicitly stated otherwise.

## Results

### Power electrophysiological pattern associated with WMH volume

When assessing the existence of significant associations between the total volume of WMH and the electrophysiological power, we only found significant results for the population with mild-VI. The data-driven analysis found a significant cluster (CBPT, p value = 0.0120) in the frequency interval [17.50 – 38.00Hz] (see Figure 1, C) The cluster size oscillates between a minimum of 14 nodes at the beginning of the frequency range and 19 at the end of that frequency range (see Figur, D), reaching its maximum extension at 26.25 Hz (442 nodes). the intensity of the correlations (see Figur, E) varied between a minimum of −0.232 and a maximum of −0.333, values that were found at 29.00 Hz. As a representative marker of the cluster, we computed its average power across all nodes and significant frequency steps. We will refer to this marker henceforth as βpow. The βpow was found to be negatively correlated (rho = −0.326, p = 0.0003, see Figur, A) with the vascular damage load across the individuals with Mild-VI. When the correlation was carried out with the entire population and with the individuals with Low-VI we did not find any significant association (see Figure 1, A). The groups with Low and Mild VI did not show significant results in βpow (see Figure 1, A & B).

**Figure 1.**
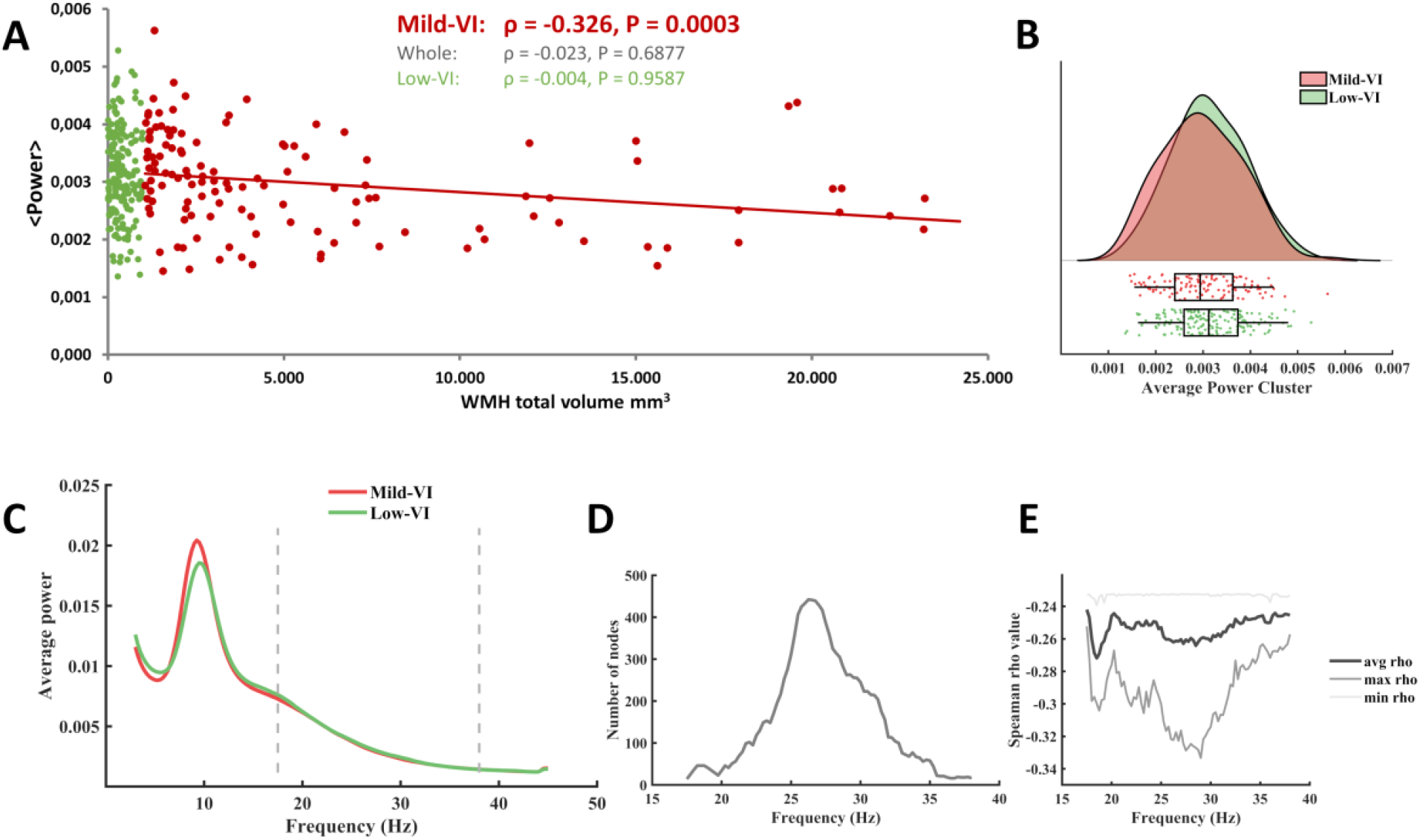
Quantitative description of the significant cluster. A, the scatter plot shows the correlations between the average power of the cluster (across all frequency steps and nodes) and the WMH total volume for the entire sample (grey), for the low VI population (green), and for the population with Mild-IV population (red). B, violin plots and boxplots graphics describing the individual values for the average power of the cluster in the significant frequency range. C, representation of the average spectral power across all significant nodes. The significant frequency interval is marked with dashes lines. D, number of grid nodes that are part of the cluster at each frequency step (maximum extension was found at 26.25 Hz). E: minimum, maximum, and average rho values across all nodes contained within the cluster at each frequency step (maximum correlation was found at 29 Hz)

**Figure 2.**
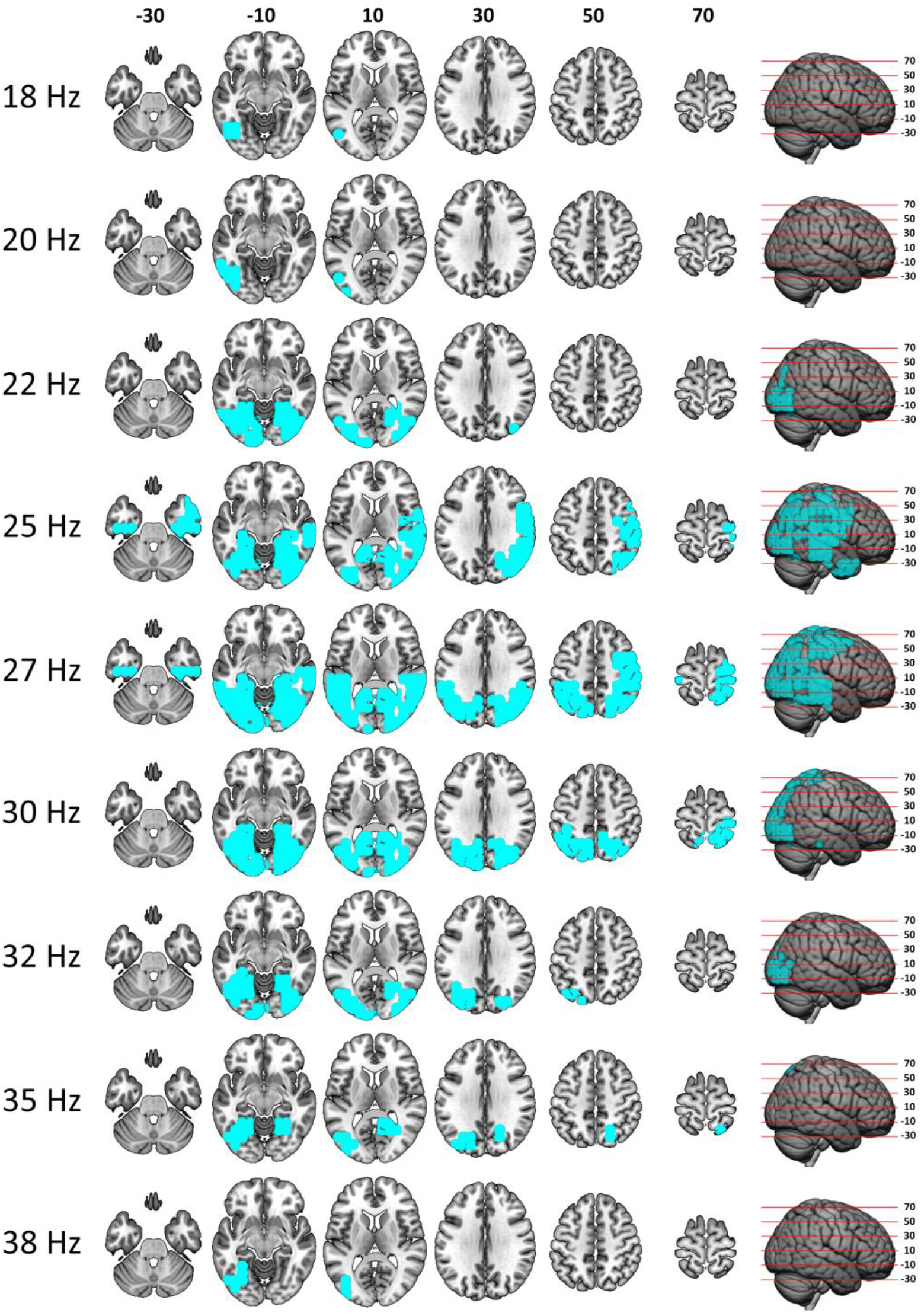
Correlation between beta spectral power and total WMH volume in the Mild-VI population. The figure shows the evolution of the cluster morphology (highlighted in cyan) through the different frequency steps. Axial slices of the brain are defined in MNI coordinates.

This β cluster mainly comprises posterior regions of the brain (see Figure and Table 2). The result indicated that an increase in vascular lesion volume appeared to be associated with a decrease in power at each frequency step of the interval in the corresponding brain regions shown in the Figure.

**Table 1.**
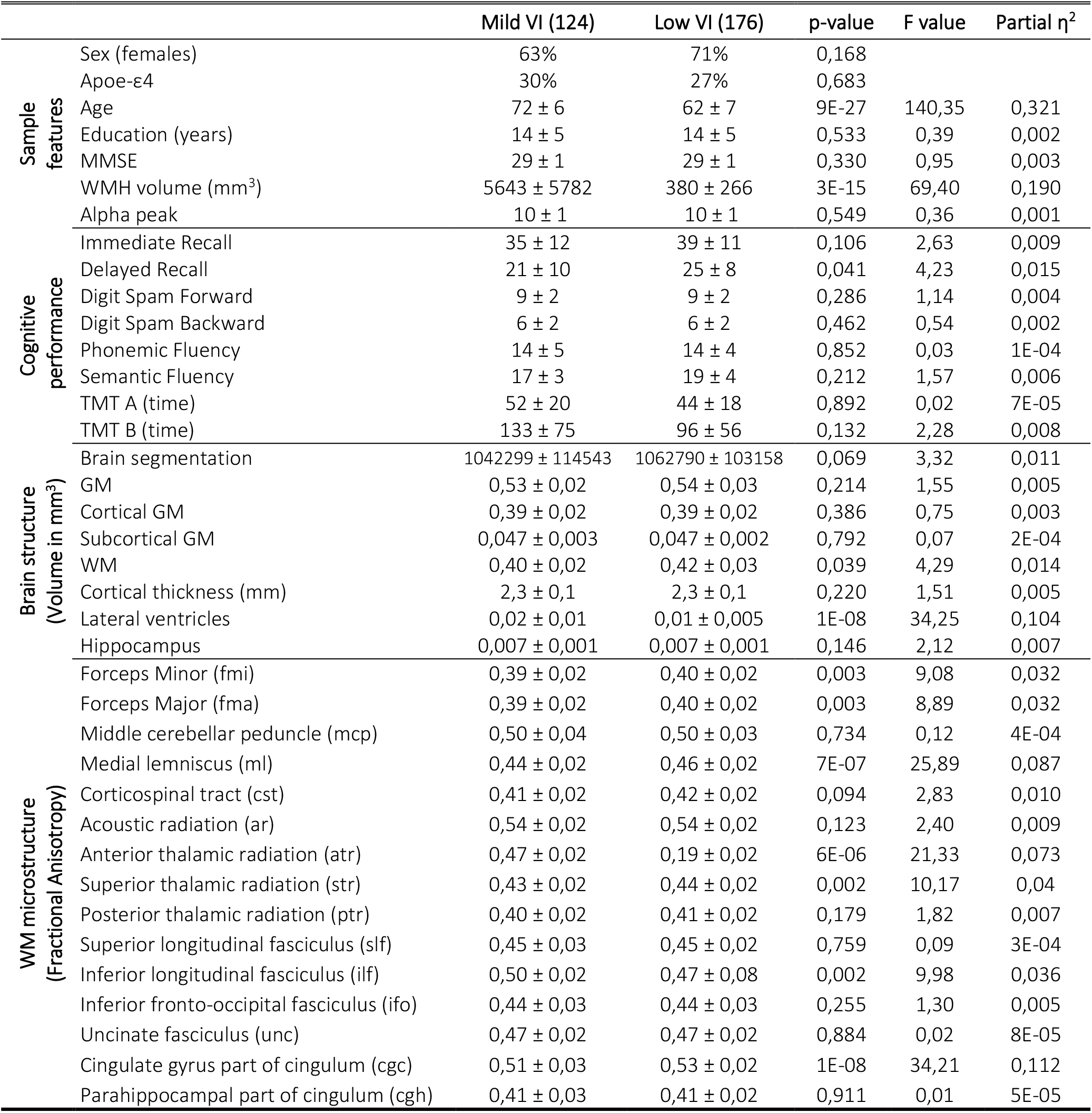
Note: Values are presented as mean ± SD. MMSE = Mini-Mental State Examination, TMT-A = Trail-Making Test part A, TMT-B = Trail-Making Test part B, GM = grey matter, WM = white matter. Each tract is the result of summing the values of that tract in both hemispheres (left and right), except Forceps major and minor and middle cerebellar peduncle because there are only one in the original atlas. p-values and F scores correspond with between-groups ANCOVA test with age and total brain volume segmentation as covariates. For age and brain segmentation volume we used between-groups ANCOVA test excluding the correspondent as covariate. For sex and ApoE-ε4 we used Fisher exact test. η2 account for partial-Eta squared for all comparisons but for the case of age and brain segmentation, where the values correspond to η2 values. Alpha peak responds for the individual alpha peak frequency obtained in the occipital-parietal region of the brain.

**Table 2.** Cluster β description (ROIs)

| ROI | % of cluster | % of ROI | Avg rho value |
| --- | --- | --- | --- |
| R Middle temporal gyrus (rMTG) | 6,79 | 81,08 | -0,259 |
| R Postcentral gyrus (rPosCG) | 6,11 | 81,82 | -0,249 |
| R Precentral gyrus (rPreCG) | 4,75 | 77,78 | -0,255 |
| L Middle Occipital lobe (lMOccL) | 4,75 | 72,41 | -0,259 |
| R Superior Temporal gyrus (rSTG) | 4,30 | 70,37 | -0,253 |
| R Angular gyrus (rAng) | 4,07 | 100,00 | -0,265 |
| R Middle Occipital lobe (rMOccL) | 3,85 | 100,00 | -0,280 |
| R Fusiform gyrus (rFusiG) | 3,85 | 89,47 | -0,277 |
| R Superior Parietal gyrus (rSPG) | 3,85 | 94,44 | -0,269 |
| L Inferior Temporal gyrus (lITG) | 3,62 | 66,67 | -0,251 |
| R Lingual gyrus (rLingual) | 3,39 | 83,33 | -0,267 |
| L Inferior Parietal gyrus (lIPG) | 3,39 | 83,33 | -0,274 |
| R Inferior Temporal gyrus (rITG) | 3,39 | 57,69 | -0,266 |
| L Calcarine fissure and surrounding cortex (lCalc) | 3,17 | 70,00 | -0,256 |
| L Precuneus (lPrecu) | 2,71 | 42,86 | -0,254 |
| L Fusiform gyrus (lFusiG) | 2,49 | 73,33 | -0,289 |
| L Superior Parietal gyrus (lSPG) | 2,49 | 68,75 | -0,260 |
| L Middle temporal gyrus (lMTG) | 2,49 | 25,00 | -0,255 |
| L Lingual gyrus (lLingual) | 2,26 | 71,43 | -0,265 |
| R Superior Occipital lobe (rSOccL) | 2,26 | 100,00 | -0,265 |
| R Inferior Parietal gyrus (rIPG) | 2,26 | 90,91 | -0,249 |
| R Supramarginal gyrus (rSMG) | 2,26 | 100,00 | -0,261 |
| L Angular gyrus (lAng) | 2,04 | 100,00 | -0,276 |
| R Precuneus (rPrecu) | 2,04 | 42,86 | -0,270 |
| R Middle Frontal gyrus (rMFG) | 1,81 | 20,00 | -0,252 |
| R Calcarine fissure and surrounding cortex (rCalc) | 1,81 | 66,67 | -0,270 |
| R Cuneus (rCu) | 1,81 | 61,54 | -0,263 |
| R Hippocampus (rHip) | 1,13 | 71,43 | -0,265 |
| L Superior Occipital lobe (lSOccL) | 1,13 | 45,45 | -0,251 |
| L Inferior occipital lobe (lIOccL) | 1,13 | 62,50 | -0,280 |
| R Inferior occipital lobe (rIOccL) | 1,13 | 100,00 | -0,282 |
| L Precentral gyrus (lPreCG) | 1,13 | 14,71 | -0,245 |

**% ROI** = percentage of the ROI within the cluster. **% Clus** = percentage of the cluster within the ROI. **Avg rho value** = average rho value obtained for the Spearman correlation between WMH load and $\beta$ power across all nodes involved within the corresponding ROI. ROIs were ordered based on their cluster size percentage. Only ROIs that contained more than 1% of the cluster are listed. r/l=right/left.

### Brain structure and cognition linked to the electrophysiological βpow marker

Finally, we used βpow to subsequent correlation analyses with cognitive performance and brain structure (see Table 3) in the entire population and for each sample based on the VI load. The results of these analyses showed a clear relationship between higher βpow and better structural brain integrity. In the entire population, βpow was found to be negatively correlated with age and positively correlated with the individual alpha central frequency. In the entire population and especially in the population with medium damage, a strong relationship was found between white matter structural integrity markers and βpow. The only conflicting results consisted of negative correlations between brain and gray matter volume with βpow, indicating that higher brain volume appeared to be associated with lower βpow.

**Table 3.**
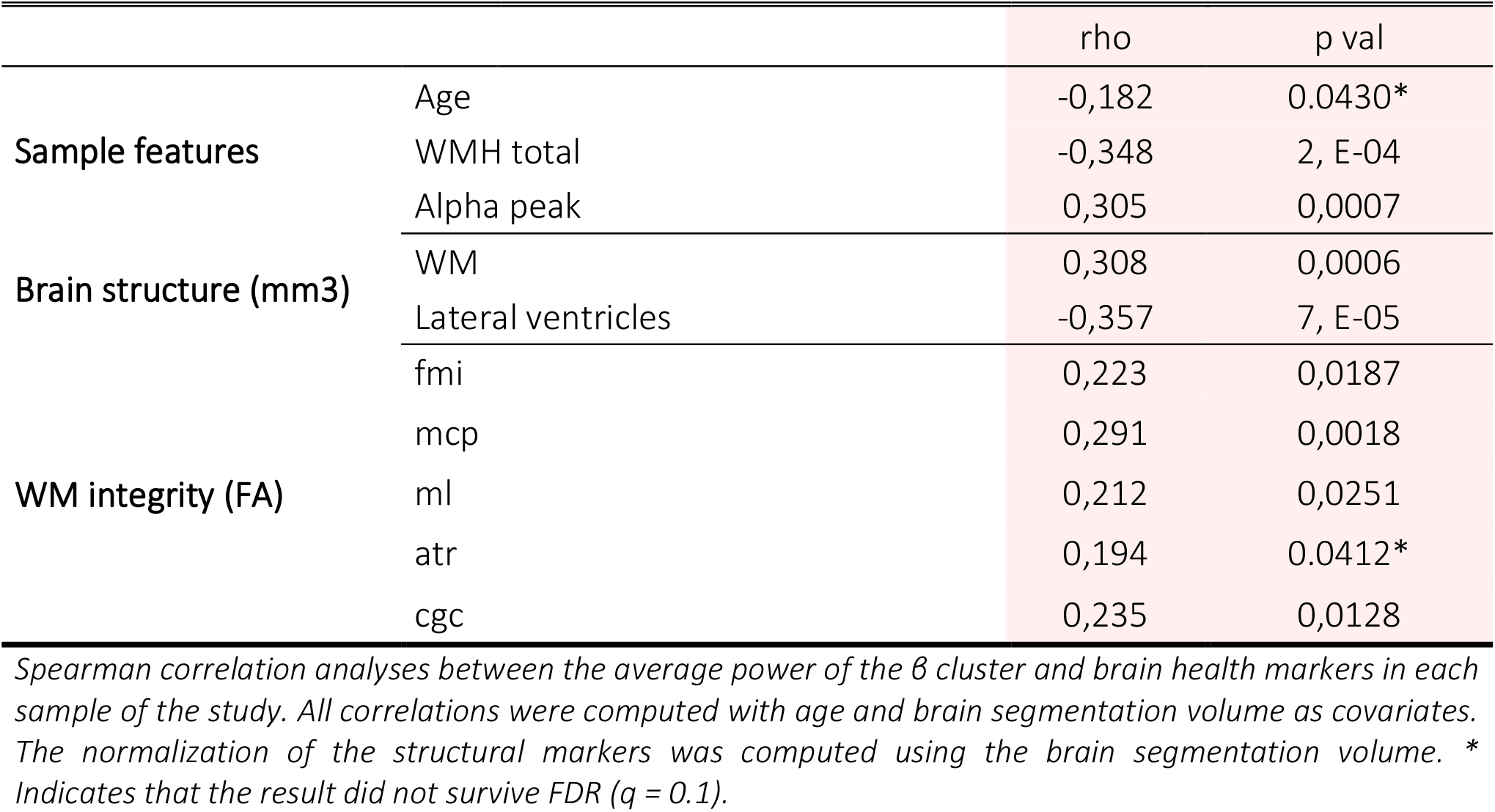
significant correlation between β power marker and brain health in the mild-VI group.

## Discussion

Our experimental study aimed to identify resting-state MEG power spectra patterns linked to cerebrovascular damage (WMH) and their impact on cognitive performance and brain structure integrity in cognitively healthy aging individuals. We hypothesized a “slowness” pattern with increased δ and θ power and decreased β power correlating with vascular damage severity. However, our findings partially supported this, revealing a significant association between lower β power and WMH volume but not the expected increase in δ and θ power.

Our study’s hypothesis, that lower β-band power correlates with increased WMH volume, was partially confirmed. Although we did not observe the expected increases in δ and θ power, the negative association between β power and WMH volume is a significant finding. This is particularly relevant given the scarcity of MEG studies examining WMH in cognitively healthy older adults. Our predictions were grounded in EEG research with broader samples that demonstrated slow-band power increases—δ and θ—in patients with cognitive impairments and cerebrovascular damage when compared to controls (Al-Qazzaz, Ali, et al., 2017; Al-Qazzaz, Hamid Bin Mohd Ali, et al., 2017; Babiloni, Binetti, et al., 2004; Moretti et al., 2004; Neto et al., 2015; Schreiter Gasser et al., 2008; van Straaten et al., 2012; Wu et al., 2014). Moreover, such increases were more marked in VCI than AD, despite similar cognitive deficits (Babiloni, Binetti, et al., 2004; Gawel et al., 2009; Moretti et al., 2007; Quandt et al., 2020; Sheorajpanday et al., 2013b).

The lack of δ and θ power augmentation in our study might be attributed to the cognitive health of our participants (MMSE ≥ 26), suggesting that the “slowness” pattern may not be present in the absence of cognitive symptoms. However, the observed decrease in β-power is consistent with existing literature on VCI and may be an early biomarker of vascular damage, preceding cognitive decline. This association was particularly strong among participants with higher levels of vascular damage (Mild-VI), and it aligned with structural brain changes such as ventricular enlargement, white matter volume reduction, and fractional anisotropy loss in significant tracts (fmi, mcp, ml, atr, cgc). Notably, β-power did not correlate with current cognitive performance but may forecast future impairment.

Our results extend previous findings by demonstrating a relationship between decreased β-band power and increased WMH volume in a cognitively healthy aging population. The study advances our understanding of the electrophysiological changes associated with subclinical cerebrovascular damage. It suggests that MEG could be a valuable tool for early detection of cerebral vascular changes that precede cognitive decline. Nevertheless, further research is needed to explore the temporal dynamics of these electrophysiological changes and their predictive value for cognitive deterioration. Subsequent studies should consider longitudinal designs to track these electrophysiological markers over time and validate their prognostic significance.

### Limitations

This study’s findings must be interpreted within the context of its limitations. Primarily, the use of MEG to investigate electrophysiological signatures associated with vascular damage is less common compared to EEG, which may limit the comparability of our results with the existing body of research. Additionally, the cross-sectional design precludes the establishment of causality between observed electrophysiological patterns and cerebrovascular damage progression. Our sample was limited to cognitively healthy elderly participants; thus our results may not generalize to individuals with existing cognitive impairments or younger cohorts. Furthermore, while we aimed to control for confounding variables such as age and brain volume, there may be other unmeasured factors, such as lifestyle or genetic predispositions, that could influence both brain health and electrophysiological signals. The study also did not account for the full spectrum of vascular pathology, focusing only on WMH volume rather than the diversity of vascular changes that may occur in aging brains. Finally, although we employed rigorous statistical methods to analyze our data, the risk of type II errors persists due to the complex nature of the signals and the stringent correction for multiple comparisons. Future longitudinal studies with larger and more diverse populations are needed to validate our findings and further elucidate the relationship between electrophysiological changes and vascular brain damage.

## Conclusions

we observed a significant decrease in β band power related to vascular damage severity but no increase in slow band power. The βpow marker correlated with structural brain changes but not with cognitive performance, indicating its potential as an early indicator of vascular damage risk in cognitively healthy older adults.

## Data Availability

The data that support the findings of this study are available from the corresponding author, upon request. All the algorithms used in the present article are reported in the Methods section.

